# Neuroplasticity-Based Treatment Prioritization in Major Depressive Disorder: A Two-Cohort Study

**DOI:** 10.64898/2026.10.06.26364759

**Authors:** Talia Cohen Solal, Revital Amiaz, Moria Maman, Ziv Kassner, Guy Keller, Dror Ben Ami, Malgorzata Jaremko, Etay Aloni, Orit Goldman, Maurizio Fava, Daphna Laifenfeld

## Abstract

**Background:** Antidepressant selection in major depressive disorder remains trial-and-error, and no biomarker identifies which drug will work for an individual patient. Because neuroplasticity is a convergent mechanism of antidepressant response, we tested whether it could serve as a drug-specific biomarker in patient-derived neurons.

**Methods:** We developed the Neuroplasticity Score (NPS), integrating structural and synaptic features from high-content imaging of patient-derived cortical neurons, and validated it against graded brain-derived neurotrophic factor (BDNF) exposure in neurons from 16 healthy donors. NPS was then applied unmodified to 75 antidepressant treatment episodes from 44 patients across two independent retrospective cohorts: 40 episodes from 20 STAR*D participants (Cohort 1; USA multi-site) and 35 episodes from 24 participants at Sheba Medical Center and Geha Mental Health Center (Cohort 2; Israel).

**Results:** NPS increased with escalating BDNF dose (Spearman ρ=0.83; P<.001). Response rate rose monotonically across NPS tertiles in both cohorts (trend P=.001 and P<.001). Across all 75 episodes, 84.6% of episodes in the highest NPS tertile responded, compared to 48.0% overall. NPS discriminated responding from non-responding episodes with an area under the receiver operating characteristic curve of 0.81 in both cohorts. The pooled odds ratio per 1-SD increase was 6.05 (95% CI, 2.80-13.05; I²=26%). Within patients contributing both a responding and a non-responding episode, the highest-scoring drug was the one that worked in 86% of patients (P<.001).

**Conclusions:** Higher antidepressant-induced neuroplasticity in a patient’s own neurons corresponded to a higher likelihood of response to that specific drug across two independent cohorts.

## Introduction

Major depressive disorder (MDD) affects over 300 million people and remains a leading cause of disability worldwide (1). Despite the availability of multiple antidepressant classes, treatment selection continues to rely on a trial-and-error process in which most patients are prescribed several ineffective drugs before achieving meaningful improvement (2,3). Delayed or inadequate response prolongs illness course and worsens functional outcomes (2,4). A tool that identifies which antidepressant will work for a given patient is therefore needed.

To date, research has not been able to identify sufficiently accurate and/or practical biomarkers for antidepressant response. Inflammatory cytokines such as IL-6 and TNF-α, neuroimmune and transcriptomic signatures (5,6), neurotrophins including BDNF (7), and metabolomic markers (8) show associations with outcome, but reproducibility has been limited and effect sizes modest. Neurophysiologic and neuroimaging biomarkers offer higher predictive potential, including rostral anterior cingulate theta activity predicting symptom improvement irrespective of treatment arm (9), reward-processing engagement predicting bupropion benefit (10), and fMRI-based connectivity subtypes predicting treatment response (11), but are costly and not widely accessible.

Across mechanistically distinct treatment modalities, impaired neuroplasticity has emerged as a convergent biological feature of MDD. Depression is characterized by reduced synaptic density, diminished dendritic complexity, and maladaptive stress-induced structural remodeling (12,13), and restoration of plasticity is increasingly regarded as a common mechanism of antidepressant efficacy (14). In patients, post-mortem studies show fewer prefrontal synapses (15), synaptic-density PET shows lower density in proportion to symptom severity (16), and non-invasive measures of LTP-like cortical plasticity are impaired (17,18); conversely, citalopram increases hippocampal grey matter (19), antidepressant-treated patients show more hippocampal neural progenitor cells (20), and ketamine enhances LTP-like plasticity (21) and prefrontal connectivity in responders (22). Chronic antidepressant exposure enhances synaptogenesis and neurotrophic signaling (23), disruption of plasticity-related pathways abolishes antidepressant efficacy (24), and the rapid clinical effects of ketamine are accompanied by BDNF-dependent synaptogenesis within hours (12,25,26). A patient-specific, functional measure of antidepressant-induced neuroplasticity could therefore address a central limitation of existing biomarker approaches.

Advances in induced pluripotent stem cell (iPSC) technology allow generation of functional cortical neurons from peripheral blood. iPSC-derived neurons retain the donor’s genetic background and capture patient-level variation in neuronal signaling and synaptic function, supporting their use as a precision psychiatry platform (27–30).

Here we developed a high-content imaging assay that quantifies drug-induced neuroplasticity in patient-derived cortical neurons, integrating structural and synaptic features into a composite Neuroplasticity Score (NPS) with dose sensitivity to graded plasticity stimulation, and evaluated across two independent cohorts whether this cellular phenotype tracks differential clinical antidepressant response, including within patients receiving multiple medications. To our knowledge, this is the first such evaluation.

## Methods and Materials

### Study population

We analyzed two independent, geographically distinct clinical cohorts. Cohort 1 comprised 40 antidepressant treatment episodes from 20 STAR*D participants (USA, multi-site), assayed from lymphoblastoid cell lines (LCLs); response was defined as ≥50% reduction in QIDS score. Cohort 2 comprised 35 treatment episodes from 24 patients treated at Sheba Medical Center and Geha Mental Health Center (Israel), whose blood was collected under IRB-approved protocols 7038-20-SMC (Sheba) and 0024-23-GEH (Geha); response was determined by clinician-rated HAMD (n=23 episodes) or by treating-clinician judgement (n=12). Together, the cohorts comprised 75 episodes from 44 patients (Figure 1). Sixteen de-identified healthy-donor PBMC specimens (Precision for Medicine) were used for the BDNF dose-response experiments. All Cohort 2 participants provided written informed consent; STAR*D participants consented under the parent study, and LCLs and clinical data were obtained under NIMH data-use terms; healthy-donor specimens were collected with donor consent by the supplier. The study was conducted in accordance with the Declaration of Helsinki.

**Figure 1.**
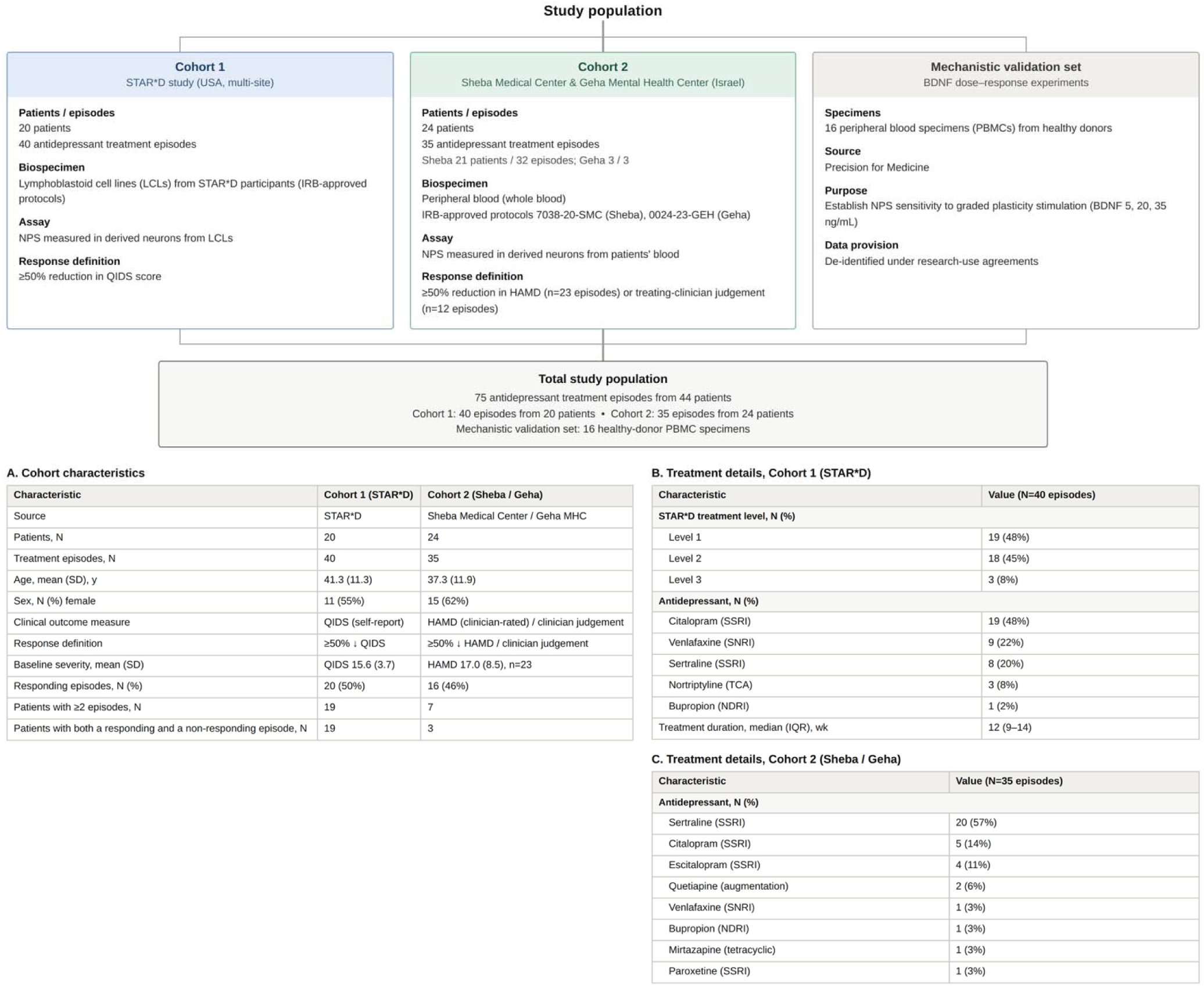
Study population and cohort characteristics. The upper schematic shows the three specimen sets contributing to the study, with the biospecimen, assay and response definition applied to each: Cohort 1 (STAR*D, USA multi-site), Cohort 2 (Sheba Medical Center and Geha Mental Health Center, Israel), and a mechanistic validation set of healthy-donor specimens used for the BDNF dose–response experiments. The two clinical cohorts together contributed 75 antidepressant treatment episodes from 44 patients. **[A]** Demographic and clinical characteristics of both clinical cohorts; patients contributing both a responding and a non-responding episode are those eligible for the within-patient analyses. **[B]** Treatment details for Cohort 1, including STAR*D treatment level, antidepressant received, and treatment duration. **[C]** Antidepressants received in Cohort 2. Percentages are of treatment episodes unless otherwise indicated. BDNF, brain-derived neurotrophic factor; HAMD, Hamilton Depression Rating Scale; IQR, interquartile range; IRB, institutional review board; LCL, lymphoblastoid cell line; MHC, Mental Health Center; NDRI, norepinephrine–dopamine reuptake inhibitor; NPS, Neuroplasticity Score; PBMC, peripheral blood mononuclear cell; QIDS, Quick Inventory of Depressive Symptomatology; SNRI, serotonin–norepinephrine reuptake inhibitor; SSRI, selective serotonin reuptake inhibitor; STAR*D, Sequenced Treatment Alternatives to Relieve Depression; TCA, tricyclic antidepressant.

### Pre-specification and blinding

NPS is a deterministic algorithm (feature extraction, normalization, aggregation and distance computation) that was fixed before linkage to clinical outcomes and not subsequently adjusted. All assays and NPS computation were performed blind to BDNF dose and clinical data.

### iPSC reprogramming

iPSCs were reprogrammed from PBMCs (BDNF validation set and Cohort 2) or from LCLs (Cohort 1) using the CytoTune-iPS 2.0 Sendai reprogramming kit (Thermo Fisher), maintained in mTeSR Plus (STEMCELL Technologies) and confirmed mycoplasma-free; the full protocol is provided in the Supplement.

### Cortical neuron differentiation

Excitatory cortical neurons were generated by doxycycline-inducible NGN2 expression as previously described with modifications (30): puromycin-selected iPSCs were seeded on Matrigel-coated 96-well plates and differentiated in sequential induction media (Table S1; full protocol in the Supplement). All selected patient-derived LCLs/PBMCs were successfully reprogrammed and differentiated into cultures enriched for excitatory cortical neurons.

### Compound/vehicle exposure

Compound concentrations were set within or near the therapeutic plasma reference range of each drug (31) and bounded above by toxicity thresholds determined empirically in our iPSC-derived neuronal cultures, which tolerate lower concentrations than the cell lines, neural progenitors and organoids used in previously published in vitro exposures (32–40). Media containing 2× compound (concentrations in Table S2) or vehicle (DMSO) was applied as a 50% exchange on days 13, 15 and 17 of differentiation, and plates were fixed in 2% paraformaldehyde on day 19.

### Imaging and quantification of the neuroplasticity score (NPS)

Fixed neurons were immunostained for GFP, PSD-95 and Synapsin1 and imaged on a Celldiscoverer 7 high-content platform (Zeiss). A standardized pipeline performed illumination correction, segmentation, and extraction of bio-objects (process, soma, junctions, spines). Bio-features capturing dendritic morphology, neurite density, and Synapsin1/PSD-95 puncta density and colocalization were z-scored, aggregated across tiles to condition-level feature vectors, and NPS computed as the Euclidean distance between treatment and matched within-plate vehicle centroids.

### Statistical analyses

NPS was analyzed as a continuous, standardized variable. For mechanistic validation, dose-dependent effects of BDNF were evaluated using within-donor repeated-measures analyses, with Spearman rank-correlation and donor-level linear-trend tests. Clinical analyses used NPS standardized within cohort, as a percentile (0–100) for tertile assignment and display and as a z-score for regression models, with effects reported per 1-SD increase, the standard deviation being the within-cohort sample standard deviation (denominator n-1), applied identically in both cohorts and in every model.

Clinical analyses were conducted at the treatment-episode level. The primary outcome was categorical response, defined as ≥50% reduction in QIDS (Cohort 1) or HAMD (Cohort 2) score or, for Cohort 2 episodes without paired HAMD scores, treating-clinician judgement; the secondary outcome was percent change in QIDS/HAMD score from baseline to end of treatment. Because both cohorts include patients contributing more than one episode, associations were evaluated using generalized estimating equations (GEE) with an independence working correlation structure and robust (sandwich) variance estimation, clustered by patient; linear and logistic GEE models were applied for continuous and binary outcomes, respectively.

The primary analysis was the gradient in observed response rate across NPS tertiles: episodes were assigned to tertiles on the within-cohort NPS percentile scale (low, ≤33.3; middle, >33.3 to ≤66.7; high, >66.7) and the monotone trend tested by the Cochran–Armitage test. The probability of response as a function of NPS percentile was estimated by logistic regression, with 95% confidence bands from patient-level cluster bootstrap resampling (500 iterations), and agreement between predicted and observed response rates (calibration) assessed across the same three tertiles. Because NPS is intended to guide drug selection, the prespecified operating characteristic of primary interest was the positive predictive value (the response rate among episodes in the upper NPS tertile, a threshold set on the score distribution rather than on outcomes), reported with the negative predictive value and exact binomial 95% confidence intervals. Secondary analyses of rank discrimination used the area under the receiver operating characteristic curve (AUC), with cluster bootstrap uncertainty. Cohort effects were combined by inverse-variance fixed-effect meta-analysis, with heterogeneity quantified by Cochran Q and I².

Within-patient analyses were restricted to patients contributing at least two treatment episodes with differing outcomes (i.e. clinical response and non-response). For each such patient, the mean NPS of responding episodes was compared with that of non-responding episodes (Wilcoxon signed-rank test), and concordance was computed as the proportion of responder/non-responder episode pairs from the same patient in which the responding episode carried the higher NPS. As a negative control for drug specificity, each episode’s NPS was re-paired with the outcome of a different drug from the same patient (within-patient derangement, 3000 iterations, pooled across cohorts) and the AUC recomputed. The partition of NPS variance between and within patients was quantified by the one-way intraclass correlation among patients contributing two or more episodes, and the variance attributable to drug identity by one-way analysis of variance. Given the small number of participants, inference was additionally based on exact permutation tests in which outcomes were permuted within patient, preserving the clustered data structure.

Analyses used Python 3.10 (statsmodels 0.14, pandas 2.1, scikit-learn 1.3, NumPy 1.26; matplotlib 3.8 for figures). Two-sided significance was set at α = .05.

## Results

### Generation of patient-derived cortical neurons

All samples were successfully reprogrammed into iPSCs and differentiated into cortical neurons using the NGN2-overexpression pipeline, showing the expected hiPSC and neuronal morphology and marker expression (Figure S1), with consistent feature extraction across donors and plates.

### Biological validation of the neuroplasticity score (NPS)

To establish NPS as a neuroplasticity measure, neurons were exposed to graded concentrations of the neuroplastogen BDNF (5, 20, 35 ng/mL). NPS increased monotonically with BDNF concentration (Spearman ρ = 0.83; P < .001), with mean values of 0.32, 2.31 and 2.90 respectively (Figure 2A) and consistently positive within-donor slopes (mean, 0.086 NPS units per ng/mL; P < .001). The increase was accompanied by greater neuritic complexity and synaptic feature density (Figure 2B), confirmed by independent manual quantification (Figure 2C, 2D). Figure S2 demonstrates that technical reproducibility across independent differentiations and staining runs was high (coefficient of variation <10%) and shows representative images.

**Figure 2.**
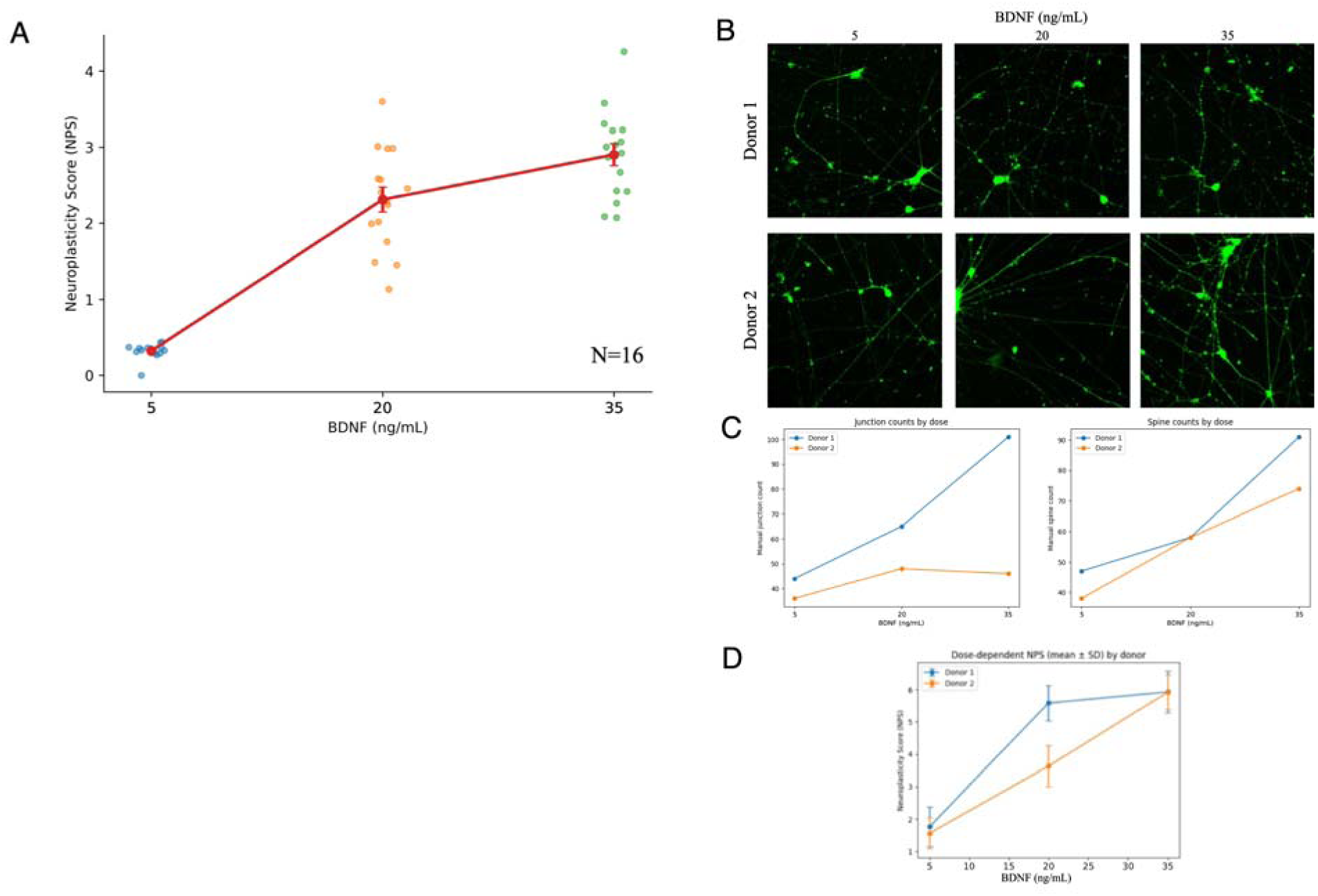
NPS reflects dose-dependent neuroplasticity in donor-derived neurons. **[A]** Donor-level NPS values are shown at 5, 20, and 35 ng/mL BDNF with jitter for visibility. Overlaid points and line represent mean ± SEM across donors. A strong monotonic association between BDNF concentration and NPS was observed (Spearman ρ = 0.83; P < .001). **[B]** Representative fluorescence images of donor-derived neurons exposed to increasing concentrations of BDNF (5, 20, 35 ng/mL), illustrating increased neuritic complexity and network connectivity at higher doses. **[C]** Manual quantification of dendritic spines and junctions in two donor lines demonstrates dose-dependent increases in neuroplasticity features, with donors shown individually to illustrate inter-donor variability. **[D]** The increase in Neuroplasticity Scores (NPS) with BDNF concentration across donor lines (mean ± SD) recapitulates manual feature quantification, with positive within-donor dose-response slopes. Manual feature quantification was performed independently and was not used in NPS derivation.

### Association of NPS with Clinical Response

Across all 75 treatment episodes contributed by 44 patients, the likelihood of responding to a given antidepressant rose monotonically with that drug’s NPS in the patient’s own neurons, in both cohorts. In Cohort 1, response occurred in 23.1% of episodes in the lowest NPS tertile (3/13), 38.5% in the middle (5/13), and 85.7% in the highest (12/14) (Cochran-Armitage trend P=.001); in Cohort 2 the corresponding rates were 9.1% (1/11), 41.7% (5/12), and 83.3% (10/12) (trend P<.001). Pooling both cohorts, response occurred in 16.7% of episodes in the lowest tertile (4/24), 40.0% in the middle (10/25), and 84.6% in the highest (22/26) (trend P<.001) (Figure 3B). Modeled as a continuous function of NPS percentile, the estimated probability of response increased steadily across the observed range in both cohorts, with closely overlapping curves (Figure 3A). In generalized estimating equation logistic models, the odds ratio per 1-SD increase in NPS was 9.97 in Cohort 1 (95% CI, 3.18-31.20; P<.001) and 3.99 in Cohort 2 (95% CI, 1.41-11.30; P=.009); given the small number of participants, both were confirmed by exact within-patient permutation tests (P<.001 and P=.037). The association was not driven by the method of outcome assessment: restricted to the episodes with clinician-rated HAMD scores, AUC was 0.80 (95% CI, 0.57-0.97; OR per 1-SD, 3.97; P=.023), and among episodes assessed by treating-clinician judgement it was 0.83. Fixed-effect meta-analysis gave a pooled odds ratio of 6.05 per 1-SD increase (95% CI, 2.80-13.05; P<.001), with consistent estimates across cohorts (I²=26%; Q-test P=.25). Adjustment for baseline symptom severity did not change these conclusions (adjusted OR, 10.02 in Cohort 1 [95% CI, 3.30-30.38] and 4.04 in Cohort 2 [95% CI, 1.20-13.66; n=23 episodes with a paired baseline score]), and baseline severity itself was not associated with response in either cohort.

**Figure 3.**
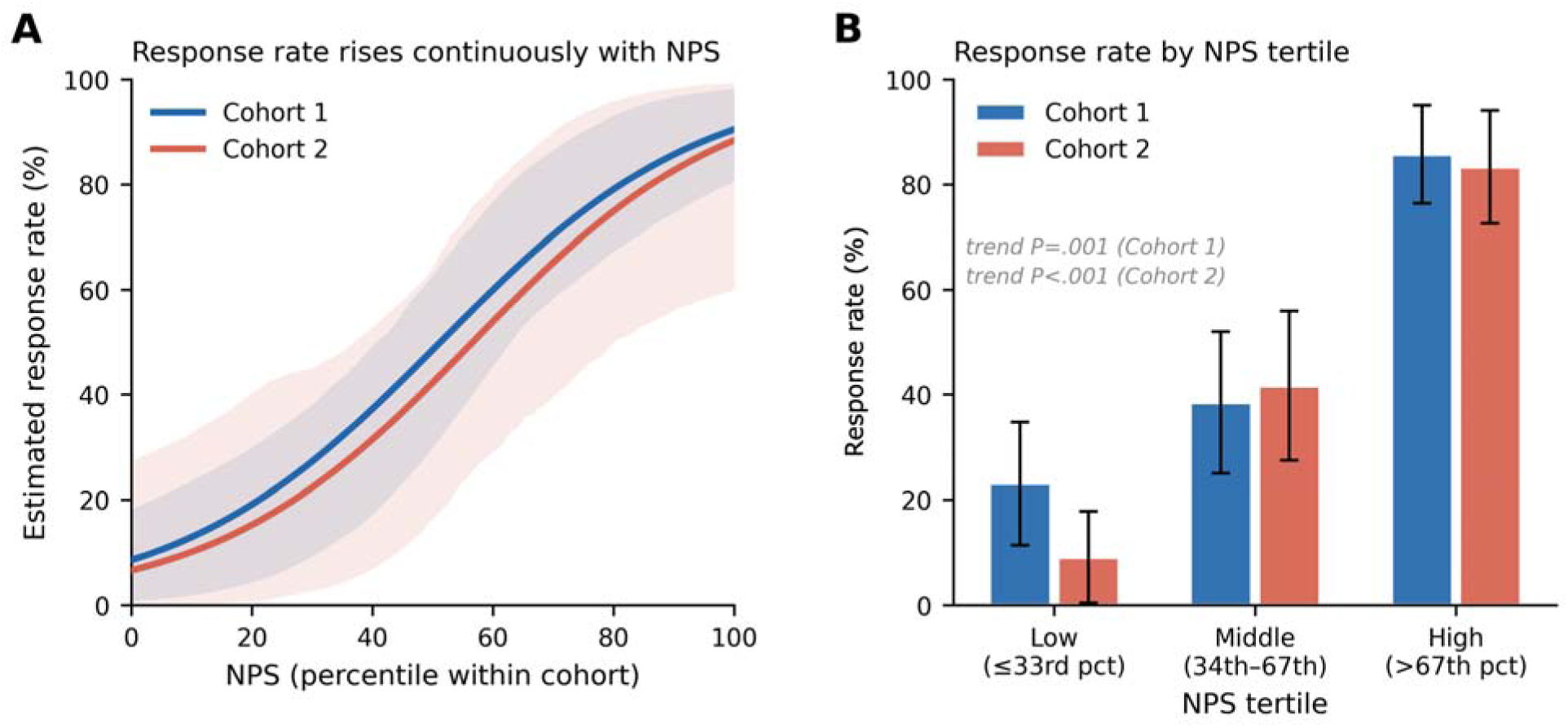
Response Rate as a Function of the NPS, Across 75 Treatment Episodes. NPS is expressed as a within-cohort percentile; this transformation is monotone and leaves rank-based statistics unchanged. [A] Estimated response rate as a continuous function of NPS percentile in each cohort (logistic fit; shaded areas are 95% patient-cluster bootstrap confidence intervals). [B] Observed response rate by NPS tertile in each cohort; response was defined as a ≥50% reduction in Quick Inventory of Depressive Symptomatology (QIDS) score in Cohort 1 and in Hamilton Depression Rating Scale (HAMD) score or, where unavailable, treating-clinician judgement in Cohort 2. Bars show observed response rates; error bars indicate ±1 binomial standard error. Tertiles are defined on the NPS percentile scale (low, ≤33.3; middle, >33.3 to ≤66.7; high, >66.7). Cochran-Armitage trend P=.001 (Cohort 1) and P<.001 (Cohort 2).

### NPS discriminates clinical response

Response probabilities implied by NPS corresponded closely to the rates actually observed: in Cohort 1, predicted clinical response rates of 18%, 49%, and 81% across the three NPS tertiles corresponded to observed rates of 23%, 38%, and 86%, and in Cohort 2, predicted clinical response rates of 14%, 43%, and 78% corresponded to 9%, 42%, and 83% observed rates (Figure S3). For treatment selection, the relevant comparison is that 48.0% of all 75 episodes met response criteria, whereas 84.6% of the 26 episodes in the highest NPS tertile did so (positive predictive value; 95% CI, 65.1%-95.6%), against 28.6% in the two lower tertiles, raising the probability that a given antidepressant would work from roughly one in two to roughly six in seven, an absolute increase of 36.6 percentage points. This held separately in each cohort (85.7% versus a 50.0% base rate in Cohort 1; 83.3% versus 45.7% in Cohort 2). NPS discriminated responding from non-responding episodes with an AUC of 0.810 in Cohort 1 (patient-cluster bootstrap 95% CI, 0.69-0.92), 0.812 in Cohort 2 (95% CI, 0.61-0.95), and 0.809 across all 75 episodes combined (95% CI, 0.70-0.90) (Figure 4). Positive and negative predictive values at the same threshold are given in Table 1, as is the within-patient prioritization accuracy.

**Figure 4.**
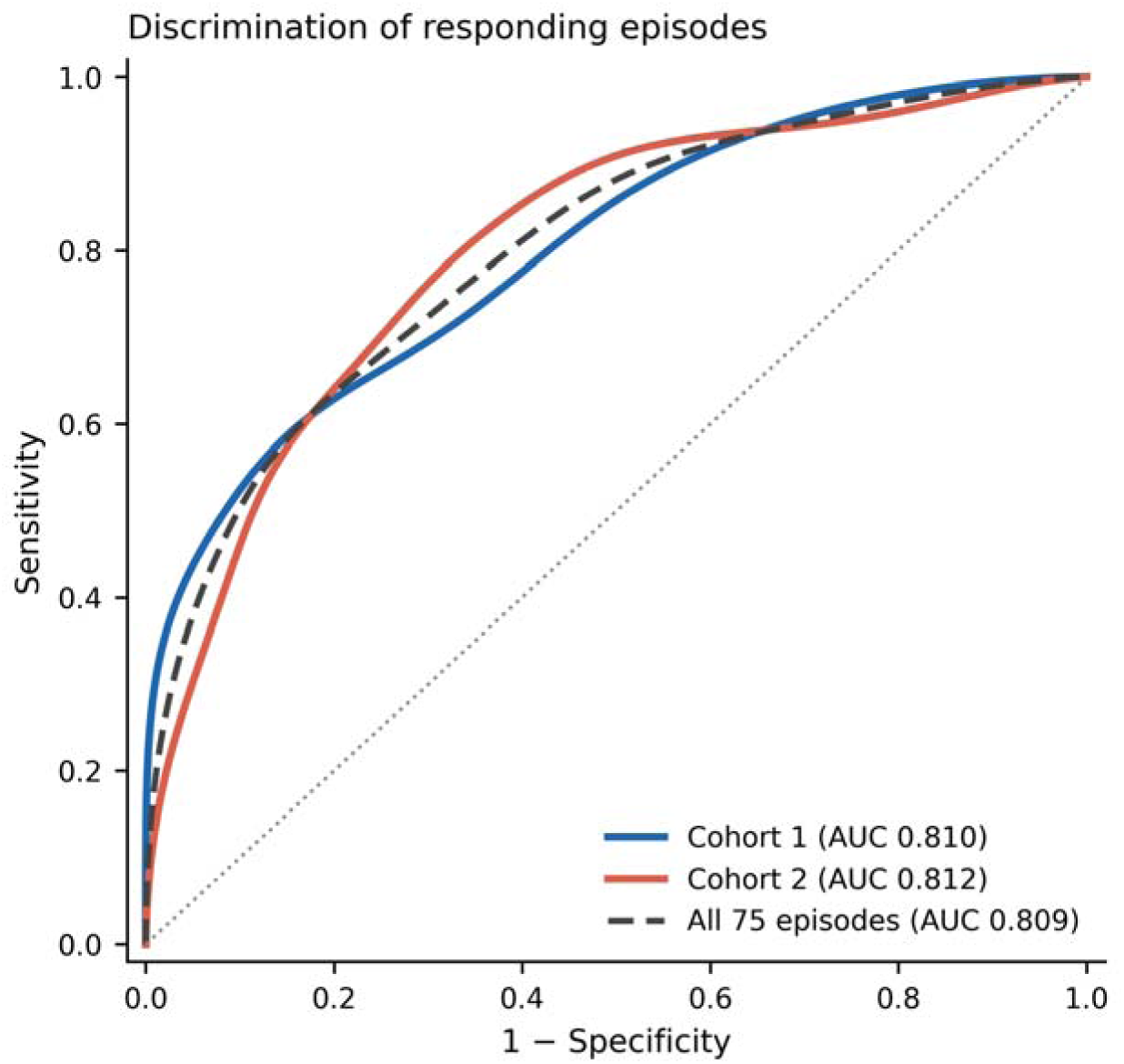
Discrimination of Responding Treatment Episodes by NPS. Receiver operating characteristic curves for discrimination of responding from non-responding treatment episodes, shown for each cohort separately and for all 75 episodes combined. NPS is expressed as a within-cohort percentile. The dotted diagonal is chance performance. AUC, area under the receiver operating characteristic curve.

**Table 1.** Performance of the Neuroplasticity Score in identifying responding antidepressant treatment episodes.

| Metric | Cohort 1 (40 episodes) | Cohort 2 (35 episodes) | All 75 episodes |
| --- | --- | --- | --- |
| **Observed response, n/N (%)** |  |  |  |
| All episodes (pre-test probability) | 20/40 (50.0) | 16/35 (45.7) | 36/75 (48.0) |
| High NPS tertile (>66.7th percentile) | 12/14 (85.7) | 10/12 (83.3) | 22/26 (84.6) |
| Two lower NPS tertiles | 8/26 (30.8) | 6/23 (26.1) | 14/49 (28.6) |
| Difference, high tertile vs all episodes | +35.7 points | +37.6 points | +36.6 points |
| **Performance at the high-tertile threshold, % (95% CI)** |  |  |  |
| Positive predictive value | 85.7 (57.2-98.2) | 83.3 (51.6-97.9) | 84.6 (65.1-95.6) |
| Negative predictive value | 69.2 (48.2-85.7) | 73.9 (51.6-89.8) | 71.4 (56.7-83.4) |
| **Discrimination across the full score range, AUC (95% CI)** |  |  |  |
| Area under the ROC curve | 0.810 (0.69-0.92) | 0.812 (0.61-0.95) | 0.809 (0.70-0.90) |
| **Within-patient prioritization, patients n/N (%)** |  |  |  |
| Accuracy (higher-NPS drug corresponds to response) | 16/19 (84.2) | 3/3 (100) | 19/22 (86.4) |

NPS tertiles are defined on the within-cohort NPS percentile scale: low, $\leq 33.3$ ; middle, $> 33.3$ to $\leq 66.7$ ; high, $> 66.7$ . The high tertile was prespecified as the operating threshold and was not derived from outcomes. Confidence intervals are exact binomial (Clopper-Pearson), other than for AUC, which uses a patient-level cluster bootstrap.

### Association of NPS with Symptom Improvement

Higher NPS was also associated with greater symptom improvement. In unadjusted GEE models, each 1-SD increase in NPS corresponded to a 21.4 percentage-point greater improvement in QIDS in Cohort 1 (95% CI, 14.5-28.3; P<.001) and a 19.1 percentage-point greater improvement in HAMD in Cohort 2 (95% CI, 0.4-37.7; P=.046; n=23 episodes with clinician HAMD scoring). After adjustment for baseline severity, age and sex (and, in Cohort 1, STAR*D treatment level), the association remained significant in Cohort 1 (β=9.3 points; 95% CI, 1.7-16.9; P=.017) and was directionally consistent in Cohort 2 (β=16.6 points; 95% CI, −2.7 to 35.9; P=.087).

### Within-Patient Treatment Prioritization

Because most patients contributed episodes on more than one antidepressant, NPS could be evaluated within individuals, with each patient serving as their own control. Among the 22 patients contributing both a responding and a non-responding episode, selecting the drug with that patient’s highest NPS identified a responding episode in 19 (86.4%; 95% CI, 65.1%-97.1%), versus 50.0% expected under random selection. The best-scoring drug that worked sat at the 67th NPS percentile on average, against the 35th percentile for the best-scoring drug that did not (Wilcoxon signed-rank P<.001) (Figure 5; per-episode detail in Figure S4). Considering every responder/non-responder episode pair drawn from the same patient, the responding episode carried the higher NPS in 88.5% of pairs (23 of 26; permutation P<.001). NPS behaved as a property of the pairing between a drug and a patient’s neurons rather than of either alone. It was not a fixed patient trait: in Cohort 1 essentially all NPS variance lay between a patient’s own drugs rather than between patients (intraclass correlation 0.00), and in Cohort 2 about one third did (ICC 0.67). Nor was it a fixed drug attribute: in Cohort 2, drug identity explained little of the variance in NPS (R²=0.10; P=.88), and among the 20 sertraline episodes NPS still separated responders from non-responders (AUC 0.81); in Cohort 1, drug identity cannot be separated from treatment level and outcome, since every sertraline episode was a Level 2 response. Re-pairing each episode’s NPS with the outcome of a different drug from the same patient reversed discrimination (AUC 0.818 vs 0.213 across the 48 episodes from these 22 patients; 3000 derangements); because most of these patients contributed two episodes, this largely mirrors the within-patient concordance above rather than constituting an independent test. NPS did not impose a fixed ranking of drugs: in 4 of 19 Cohort 1 patients the Level 1 agent (citalopram) scored higher than the switch agent, and in Cohort 2, where drugs were not given in a protocol order, NPS was unrelated to exposure order.

**Figure 5.**
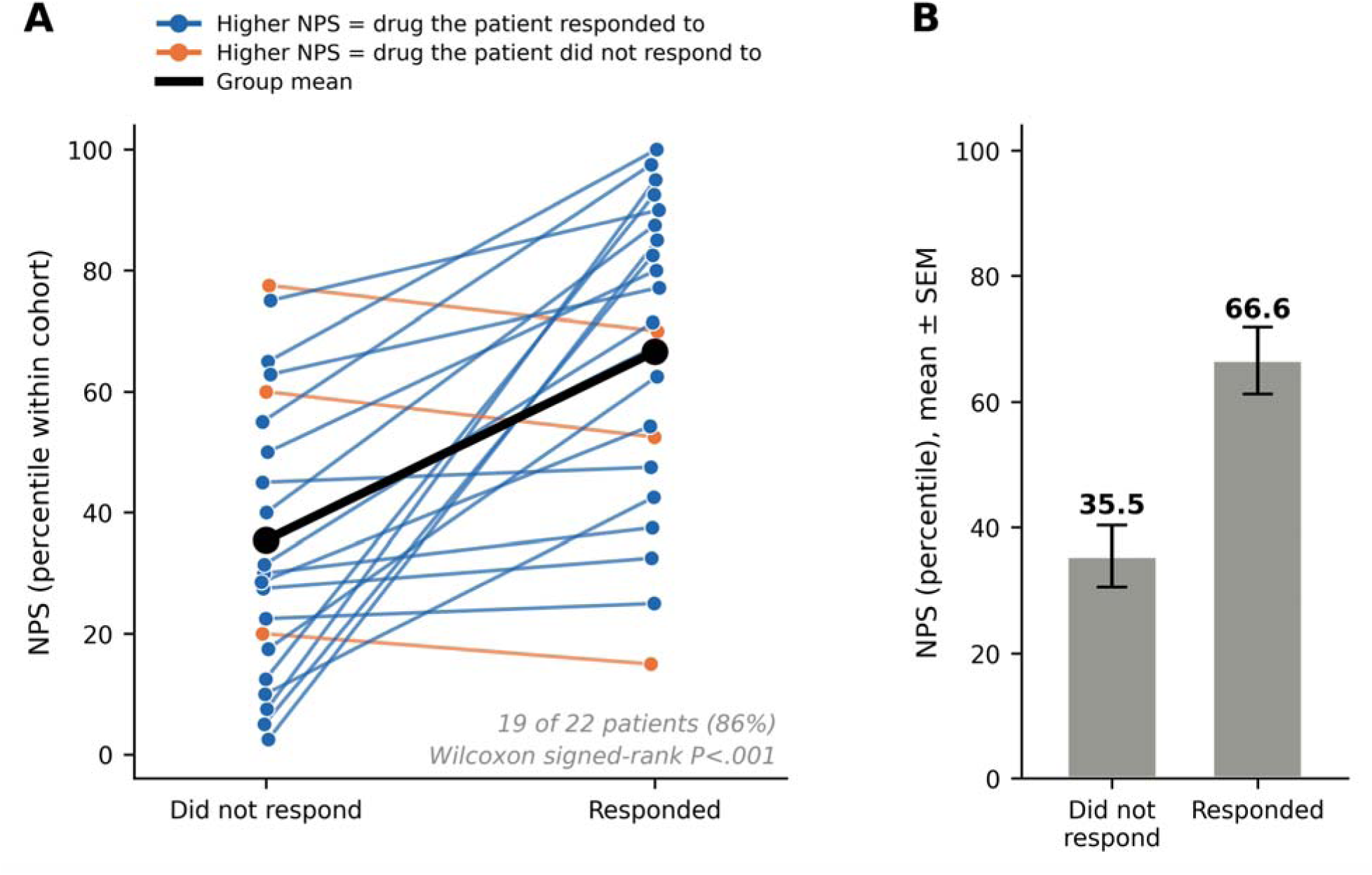
Within-Patient Correspondence Between NPS and Antidepressant Response. Analyses include the 22 patients who contributed both a responding and a non-responding treatment episode, for whom a within-patient comparison is defined; patients whose episodes shared a single outcome are not shown but are included in all other analyses. NPS is expressed as a within-cohort percentile. [A] For each patient, the highest NPS among antidepressants the patient did not respond to is joined to the highest NPS among those the patient did respond to. Blue lines indicate the 19 of 22 patients (86%) in whom the drug with the higher NPS was the one they responded to; orange lines the three in whom it was not. The black line is the group mean. [B] Group means for the same two quantities (mean ± SEM). Wilcoxon signed-rank P<.001. Every individual treatment episode for these patients is shown in Figure S4.

Because Cohort 1 patients were selected from STAR*D for having failed one treatment level and responded at a later one, response in this cohort is tied to treatment level (5% at Level 1, 89% at Level 2, 100% at Level 3), and NPS was also higher at later levels (ρ=0.43; P=.006). Within level, NPS continued to separate responding from non-responding episodes (AUC 0.83 at Level 1 and 0.81 at Level 2, although each stratum contained few episodes of one outcome). In a GEE model including treatment level, the NPS odds ratio was essentially unchanged (9.05 per 1-SD) but no longer significant (P=.073), reflecting the near-collinearity of level and outcome imposed by the selection. Cohort 2 has no such structure: response did not vary with exposure order (48%, 38% and 50% for first, second and third drugs; ρ=−0.04; P=.80), NPS was unrelated to order (ρ=−0.12; P=.48), and NPS discriminated response within each exposure position (AUC 0.80, 0.73 and 1.00). Line of therapy therefore cannot account for the association in Cohort 2.

## Discussion

The findings in this study show that the neuroplastic response of a patient’s derived neurons to a specific antidepressant is associated with their response to that drug in the clinic. Across 75 treatment episodes from 44 patients in two independent cohorts, recruited on different continents, treated under different protocols and assessed with different clinical instruments, a prespecified score, whose algorithm was frozen before linkage to any clinical data and applied without refitting, discriminated responding from non-responding depression episodes with an AUC of 0.81. Clinically, this means that while 48% of episodes overall met response criteria, 85% of those in the highest neuroplasticity score (NPS) tertile did, raising the probability that the drug under consideration would work from roughly one in two to roughly six in seven. The relationship was graded rather than dichotomous (17% response in the lowest NPS tertile versus 85% in the highest), and predicted probabilities matched observed rates. Despite differences in country, recruitment, treatment protocol, rating instrument and the absolute scale of the assay, the two cohorts produced closely consistent estimates (I²=26%; heterogeneity P=.25). Reproducibility of this kind across independent samples is uncommon among candidate antidepressant-response biomarkers, and its absence has been a principal reason such biomarkers have failed to become clinical tests (41).

### Biological Validity and Assay Performance

The biological basis for NPS rests on its integration of structural and synaptic features (dendritic morphology, neurite density, and synapsin/PSD-95 puncta density and colocalization) that are canonical correlates of synaptic strength and network connectivity (42–44). Consistent with this, NPS increased monotonically with escalating BDNF across donor lines, demonstrating sensitivity to a plasticity pathway central to antidepressant action (24,45,46) and indicating that the score operates within a biologically meaningful dynamic range.

Technical reproducibility across independent differentiations and staining runs was high relative to biological signal. While in vitro systems cannot recapitulate glial and circuit-level contributions, the consistency of dose sensitivity and of the subsequent clinical associations indicates that NPS reliably captures a cellular phenotype of neuroplasticity.

### Clinical Applicability of a Blood-Derived Cellular Assay

Antidepressant response biomarkers reported to date, such as cytokines, neurotrophins and transcriptomic signatures, reflect the systemic milieu and index central neurobiology only indirectly (6,47,48). Neuroimaging biomarkers are mechanistically closer (49) but remain constrained by cost and infrastructure (50) and have yet to show generalizable clinical utility (51,52). NPS instead interrogates patient-specific neurons directly, and its input is a standard blood draw already routine in psychiatric practice (53), with reprogramming, differentiation and imaging performed centrally. Turnaround time is therefore the principal practical constraint on deployment and should be characterized prospectively alongside clinical utility.

### Association of NPS with antidepressant response

Three features of these data support the validity and generalizability of the findings. First, the association of NPS and clinical response is graded and calibrated: response probability rose monotonically across NPS tertiles in each cohort separately (trend P=.001 and P<.001), and predicted rates matched observed rates. A spurious association would not be expected to reproduce a smooth dose–response relationship twice, in independent samples, on different assay scales. Second, the association is present within individual patients: selecting each patient’s highest-NPS drug identified a responding episode in 86.4% of patients against 50.0% expected by chance. Because the measurement itself is generated in cultured neurons, which have no capacity for such effects, the non-specific determinants of improvement (expectancy and placebo response, regression to the mean, spontaneous remission, therapeutic contact) cannot explain why NPS specifically tracks which drug worked within a patient. The within-patient comparison additionally holds stable patient-level traits constant across episodes, though it does not rule out confounds that vary with treatment order over calendar time (see Limitations). The same design excludes between-patient confounding by severity, chronicity, or comorbidity, which no between-patient biomarker can rule out. Third, the signal belongs to the pairing of a particular drug with a particular patient’s neurons, not to either alone. A score indexing only a patient’s general neuroplastic capacity could not distinguish between that patient’s drugs, yet NPS did so within patients; a score indexing only a drug’s average plastogenic effect could not distinguish between patients receiving the same drug, yet in Cohort 2 NPS separated sertraline responders from non-responders. The effect sizes obtained here are at the upper end of those reported for neuroimaging predictors (10,11), gut microbiome signatures (54), and senescence-associated secretory phenotype indices (55), and were obtained with a score that was never trained on clinical outcomes, so they are not subject to the optimism bias of a model fitted and evaluated in the same data.

These associations were observed across eight antidepressants spanning five pharmacologic classes (SSRI, SNRI, NDRI, tricyclic and tetracyclic), together with quetiapine used as an augmentation agent, consistent with NPS indexing a shared downstream neuroplastic capacity rather than class-specific pharmacodynamics. Pharmacogenetic strategies, in contrast, capture how a patient metabolizes a drug but not how the patient’s neurons respond to it, and have shown modest and variable benefit in large pragmatic trials and meta-analysis (56–58); the two approaches may prove complementary rather than competitive.

### Conceptual Implications

Current practice resolves the choice of antidepressant by sequential trial and error, with each step costing six to twelve weeks. A measurement that ranks candidate drugs for an individual before treatment begins addresses this bottleneck directly, shifting selection from empirical sequencing toward biologically informed choice for the individual patient.

### Limitations

Four limitations should be considered. First, the clinical analysis was retrospective and limited in sample size; treatment was not assigned on the basis of NPS, thus NPS association with clinical outcome cannot be determined to be causal, and clinical utility cannot be established. Second, Cohort 1 patients were selected from STAR*D for having both a failed and a successful episode; this could make the within-patient concordance easier to detect than in an unselected sample. Because each selected patient contributed one failed and one successful episode, the 50% base rate in Cohort 1 reflects the sampling design rather than response in an unselected population. Cohort 2, which was not selected on this basis, produced an essentially identical AUC (0.81) and a similar gain over its own base rate (83.3% vs 45.7%); only three of its patients contributed both a responding and a non-responding episode, so its within-patient concordance (3 of 3) is supportive but not independently conclusive. Positive and negative predictive values still depend on each cohort’s own response rate and should not be extrapolated beyond it. Third, NPS values are computed relative to a within-plate vehicle centroid and standardized within cohort, and absolute values cannot be compared across plates, cohorts, or studies without re-standardization; a shared reference stimulus with a well-characterized, dose-dependent neuroplastic effect, such as the NPS response to BDNF treatment already measured in this study, could serve as a scaling anchor to establish a scale in future work. Fourth, because Cohort 1 was selected for a failed-then-successful treatment sequence, response in that cohort is tied to STAR*D treatment level, so treatment order and NPS cannot be fully separated there; the absence of any order effect in Cohort 2, where the association replicated, argues against line of therapy as the explanation.

## Conclusions

Across 75 antidepressant treatment episodes in two independent cohorts, the neuroplastic response of a patient’s own neurons to a specific drug corresponded to whether that patient responded to that drug. A high NPS identified an antidepressant that produced clinical response in 85% of episodes against a 48% response rate overall, and within individual patients the highest-scoring drug was the one the patient responded to in 86.4% of cases. These findings support the potential utility of neuroplasticity-guided antidepressant selection within precision psychiatry.

## STAR*D data and biomaterials

Bio-samples and/or data for this publication were obtained from the NIMH Repository & Genomics Resource (supported by cooperative agreement U24 MH068457), a centralized national biorepository for genetic studies of psychiatric disorders. Data and biomaterials were obtained from the limited access datasets distributed from the NIH-supported ‘Sequenced Treatment Alternatives to Relieve Depression’ (STAR*D). STAR*D focused on non-psychotic major depressive disorder in adults seen in outpatient settings. The primary purpose of this research study was to determine which treatments work best if the first treatment with medication does not produce an acceptable response. The study was supported by NIMH Contract # N01MH90003 to the University of Texas Southwestern Medical Center. The ClinicalTrials.gov identifier is NCT00021528. This manuscript reflects the views of the authors and may not reflect the opinions or views of the STAR*D Study Investigators or the NIH.

## Acknowledgments

We thank the patients who participated in this study, and the clinical and research staff of Sheba Medical Center and Geha Mental Health Center, in particular Dr. Tom Fruchtman-Steinbok for patient recruitment, sample collection and clinical data. We thank Yura Zharkovsky and Gregory Mayzel for contributing to the research.

## Author contributions (CRediT)

TCS: Conceptualization, supervision, manuscript review and editing; RA: Clinical oversight and input, manuscript review and editing; MM: Laboratory technique ideation and implementation; ZK: Data science, including data curation, research and analysis; GK: Data science, including data curation, research and analysis; DBA: Investigation, data curation, project administration; MJ: Laboratory supervision; EA: Imaging research and analysis; OG: Supervision, research, manuscript review and editing; MF: Clinical input, manuscript review and editing; DL: Conceptualization, analysis, supervision, original draft, manuscript review and editing. All authors approved the final version of the manuscript.

## Data availability

STAR*D clinical data are available to qualified researchers through the NIMH Data Archive. De-identified episode-level NPS values and outcome data supporting these findings are available from the corresponding author upon specific request.

## Alternative Text to comply with EAA

**Figure 1** Schematic of the three specimen sets contributing to the study, with tables of cohort characteristics and the antidepressants received in each cohort.

**Figure 2** Neuroplasticity score rises with increasing BDNF dose, with representative neuronal images and dose–response feature quantification.

**Figure 3** Response rate rises with the neuroplasticity score, shown as a continuous model fit and as observed rates across NPS tertiles, in both cohorts.

**Figure 4** Receiver operating characteristic curves showing that the neuroplasticity score discriminates responding from non-responding treatment episodes, with an area under the curve of about 0.81 in each cohort.

**Figure 5** Paired plot showing that in 19 of 22 patients the antidepressant with the highest neuroplasticity score is the one the patient responded to, with group means alongside.

## References

1. World Health Organization. Depressive disorder (depression). Fact sheet. Updated September 11, 2026. Accessed October 5, 2026. https://www.who.int/news-room/fact-sheets/detail/depression

2. Rush AJ, Trivedi MH, Wisniewski SR, Nierenberg AA, Stewart JW, Warden D, et al. Acute and longer-term outcomes in depressed outpatients requiring one or several treatment steps: a STAR*D report. Am J Psychiatry. 2006;163(11):1905–1917. doi:10.1176/ajp.2006.163.11.1905

3. Gaynes BN, Warden D, Trivedi MH, Wisniewski SR, Fava M, Rush AJ. What did STAR*D teach us? Results from a large-scale, practical, clinical trial for patients with depression. Psychiatr Serv. 2009;60(11):1439–1445. doi:10.1176/ps.2009.60.11.1439

4. Oluboka OJ, Katzman MA, Habert J, McIntosh D, MacQueen GM, Milev RV, et al. Functional recovery in major depressive disorder: providing early optimal treatment for the individual patient. Int J Neuropsychopharmacol. 2018;21(2):128–144. doi:10.1093/ijnp/pyx081

5. Hodes GE, Kana V, Menard C, Merad M, Russo SJ. Neuroimmune mechanisms of depression. Nat Neurosci. 2015;18(10):1386–1393. doi:10.1038/nn.4113

6. Pisanu C, Severino G, De Toma I, Dierssen M, Fusar-Poli P, Gennarelli M, et al. Transcriptional biomarkers of response to pharmacological treatments in severe mental disorders: a systematic review. Eur Neuropsychopharmacol. 2022;55:112–157. doi:10.1016/j.euroneuro.2021.12.005

7. Sen S, Duman R, Sanacora G. Serum brain-derived neurotrophic factor, depression, and antidepressant medications: meta-analyses and implications. Biol Psychiatry. 2008;64(6):527–532. doi:10.1016/j.biopsych.2008.05.005

8. Kaddurah-Daouk R, Boyle SH, Matson W, Sharma S, Matson S, Zhu H, et al. Pretreatment metabotype as a predictor of response to sertraline or placebo in depressed outpatients: a proof of concept. Transl Psychiatry. 2011;1(7):e26. doi:10.1038/tp.2011.22

9. Pizzagalli DA, Webb CA, Dillon DG, Tenke CE, Kayser J, Goer F, et al. Pretreatment rostral anterior cingulate cortex theta activity in relation to symptom improvement in depression: a randomized clinical trial. JAMA Psychiatry. 2018;75(6):547–554. doi:10.1001/jamapsychiatry.2018.0252

10. Nguyen KP, Chin Fatt C, Treacher A, Mellema C, Cooper C, Jha MK, et al. Patterns of pretreatment reward task brain activation predict individual antidepressant response: key results from the EMBARC randomized clinical trial. Biol Psychiatry. 2022;91(6):550–560. doi:10.1016/j.biopsych.2021.09.011

11. Drysdale AT, Grosenick L, Downar J, Dunlop K, Mansouri F, Meng Y, et al. Resting-state connectivity biomarkers define neurophysiological subtypes of depression. Nat Med. 2017;23(1):28–38. doi:10.1038/nm.4246

12. Duman RS, Aghajanian GK. Synaptic dysfunction in depression: potential therapeutic targets. Science. 2012;338(6103):68–72. doi:10.1126/science.1222939

13. McEwen BS, Nasca C, Gray JD. Stress effects on neuronal structure: hippocampus, amygdala, and prefrontal cortex. Neuropsychopharmacology. 2016;41(1):3–23. doi:10.1038/npp.2015.171

14. Holmes SE, Abdallah C, Esterlis I. Imaging synaptic density in depression. Neuropsychopharmacology. 2023;48(1):186–190. doi:10.1038/s41386-022-01368-4

15. Kang HJ, Voleti B, Hajszan T, Rajkowska G, Stockmeier CA, Licznerski P, et al. Decreased expression of synapse-related genes and loss of synapses in major depressive disorder. Nat Med. 2012;18(9):1413–1417. doi:10.1038/nm.2886

16. Holmes SE, Scheinost D, Finnema SJ, Naganawa M, Davis MT, DellaGioia N, et al. Lower synaptic density is associated with depression severity and network alterations. Nat Commun. 2019;10(1):1529. doi:10.1038/s41467-019-09562-7

17. Normann C, Schmitz D, Fürmaier A, Döing C, Bach M. Long-term plasticity of visually evoked potentials in humans is altered in major depression. Biol Psychiatry. 2007;62(5):373–380. doi:10.1016/j.biopsych.2006.10.006

18. Player MJ, Taylor JL, Weickert CS, Alonzo A, Sachdev P, Martin D, et al. Neuroplasticity in depressed individuals compared with healthy controls. Neuropsychopharmacology. 2013;38(11):2101–2108. doi:10.1038/npp.2013.126

19. Arnone D, McKie S, Elliott R, Juhasz G, Thomas EJ, Downey D, et al. State-dependent changes in hippocampal grey matter in depression. Mol Psychiatry. 2013;18(12):1265–1272. doi:10.1038/mp.2012.150

20. Boldrini M, Hen R, Underwood MD, Rosoklija GB, Dwork AJ, Mann JJ, et al. Hippocampal angiogenesis and progenitor cell proliferation are increased with antidepressant use in major depression. Biol Psychiatry. 2012;72(7):562–571. doi:10.1016/j.biopsych.2012.04.024

21. Sumner RL, McMillan R, Spriggs MJ, Campbell D, Malpas G, Maxwell E, et al. Ketamine enhances visual sensory evoked potential long-term potentiation in patients with major depressive disorder. Biol Psychiatry Cogn Neurosci Neuroimaging. 2020;5(1):45–55. doi:10.1016/j.bpsc.2019.07.002

22. Abdallah CG, Averill LA, Collins KA, Geha P, Schwartz J, Averill C, et al. Ketamine treatment and global brain connectivity in major depression. Neuropsychopharmacology. 2017;42(6):1210–1219. doi:10.1038/npp.2016.186

23. Duman RS, Voleti B. Signaling pathways underlying the pathophysiology and treatment of depression: novel mechanisms for rapid-acting agents. Trends Neurosci. 2012;35(1):47–56. doi:10.1016/j.tins.2011.11.004

24. Li N, Lee B, Liu RJ, Banasr M, Dwyer JM, Iwata M, et al. mTOR-dependent synapse formation underlies the rapid antidepressant effects of NMDA antagonists. Science. 2010;329(5994):959–964. doi:10.1126/science.1190287

25. Autry AE, Adachi M, Nosyreva E, Na ES, Los MF, Cheng PF, et al. NMDA receptor blockade at rest triggers rapid behavioural antidepressant responses. Nature. 2011;475(7354):91–95. doi:10.1038/nature10130

26. Abdallah CG, De Feyter HM, Averill LA, Jiang L, Averill CL, Chowdhury GMI, et al. The effects of ketamine on prefrontal glutamate neurotransmission in healthy and depressed subjects. Neuropsychopharmacology. 2018;43(10):2154–2160. doi:10.1038/s41386-018-0136-3

27. Brennand KJ, Simone A, Jou J, Gelboin-Burkhart C, Tran N, Sangar S, et al. Modelling schizophrenia using human induced pluripotent stem cells. Nature. 2011;473(7346):221–225. doi:10.1038/nature09915

28. Stern S, Santos R, Marchetto MC, Mendes APD, Rouleau GA, Biesmans S, et al. Neurons derived from patients with bipolar disorder divide into intrinsically different sub-populations of neurons, predicting the patients’ responsiveness to lithium. Mol Psychiatry. 2018;23(6):1453–1465. doi:10.1038/mp.2016.260

29. Vadodaria KC, Ji Y, Skime M, Paquola AC, Nelson T, Hall-Flavin D, et al. Altered serotonergic circuitry in SSRI-resistant major depressive disorder patient-derived neurons. Mol Psychiatry. 2019;24(6):808–818. doi:10.1038/s41380-019-0377-5

30. Avior Y, Ron S, Kroitorou D, Albeldas C, Lerner V, Corneo B, et al. Depression patient-derived cortical neurons reveal potential biomarkers for antidepressant response. Transl Psychiatry. 2021;11(1):201. doi:10.1038/s41398-021-01319-5

31. Hiemke C, Bergemann N, Clement HW, Conca A, Deckert J, Domschke K, et al. Consensus guidelines for therapeutic drug monitoring in neuropsychopharmacology: update 2017. Pharmacopsychiatry. 2018;51(1-02):9–62. doi:10.1055/s-0043-116492

32. Then CK, Liu KH, Liao MH, Chung KH, Wang JY, Shen SC. Antidepressants, sertraline and paroxetine, increase calcium influx and induce mitochondrial damage-mediated apoptosis of astrocytes. Oncotarget. 2017;8(70):115490–115502. doi:10.18632/oncotarget.23302

33. Elsworthy RJ, Crowe JA, King MC, Dunleavy C, Fisher E, Ludlam A, et al. The effect of citalopram treatment on amyloid-β precursor protein processing and oxidative stress in human hNSC-derived neurons. Transl Psychiatry. 2022;12(1):285. doi:10.1038/s41398-022-02050-5

34. Mansvelder HD, Fagen ZM, Chang B, Mitchum R, McGehee DS. Bupropion inhibits the cellular effects of nicotine in the ventral tegmental area. Biochem Pharmacol. 2007;74(8):1283–1291. doi:10.1016/j.bcp.2007.07.034

35. Doh MS, Han DMR, Oh DH, Kim SH, Choi MR, Chai YG. Profiling of proteins regulated by venlafaxine during neural differentiation of human cells. Psychiatry Investig. 2015;12(1):81–91. doi:10.4306/pi.2015.12.1.81

36. Zhang W, Wang H, Wang X, Narayanan MV, Stavrovskaya IG, Kristal BS, Friedlander RM. Nortriptyline protects mitochondria and reduces cerebral ischemia/hypoxia injury. Stroke. 2008;39(2):455–462. doi:10.1161/STROKEAHA.107.496810

37. Zivko C, Sagar R, Xydia A, Lopez-Montes A, Mintzer J, Rosenberg PB, et al. iPSC-derived hindbrain organoids to evaluate escitalopram oxalate treatment responses targeting neuropsychiatric symptoms in Alzheimer’s disease. Mol Psychiatry. 2024;29(11):3644–3652. doi:10.1038/s41380-024-02629-y

38. Correia AS, Fraga S, Teixeira JP, Vale N. Cell model of depression: reduction of cell stress with mirtazapine. Int J Mol Sci. 2022;23(9):4942. doi:10.3390/ijms23094942

39. Hosseini K, Cediel-Ulloa A, Al-Sabri MH, Forsby A, Fredriksson R. Assessing the neurodevelopmental impact of fluoxetine, citalopram, and paroxetine on neural stem cell-derived neurons. Pharmaceuticals (Basel). 2024;17(10):1392. doi:10.3390/ph17101392

40. Burbulla LF, Zheng J, Song P, Jiang W, Johnson ME, Brundin P, Krainc D. Direct targeting of wild-type glucocerebrosidase by antipsychotic quetiapine improves pathogenic phenotypes in Parkinson’s disease models. JCI Insight. 2021;6(19):e148649. doi:10.1172/jci.insight.148649

41. Kapur S, Phillips AG, Insel TR. Why has it taken so long for biological psychiatry to develop clinical tests and what to do about it? Mol Psychiatry. 2012;17(12):1174–1179. doi:10.1038/mp.2012.105

42. Hering H, Sheng M. Dendritic spines: structure, dynamics and regulation. Nat Rev Neurosci. 2001;2(12):880–888. doi:10.1038/35104061

43. Ippolito DM, Eroglu C. Quantifying synapses: an immunocytochemistry-based assay to quantify synapse number. J Vis Exp. 2010;(45):2270. doi:10.3791/2270

44. Moda-Sava RN, Murdock MH, Parekh PK, Fetcho RN, Huang BS, Huynh TN, et al. Sustained rescue of prefrontal circuit dysfunction by antidepressant-induced spine formation. Science. 2019;364(6436):eaat8078. doi:10.1126/science.aat8078

45. Casarotto PC, Girych M, Fred SM, Kovaleva V, Moliner R, Enkavi G, et al. Antidepressant drugs act by directly binding to TRKB neurotrophin receptors. Cell. 2021;184(5):1299–1313.e19. doi:10.1016/j.cell.2021.01.034

46. Castrén E, Antila H. Neuronal plasticity and neurotrophic factors in drug responses. Mol Psychiatry. 2017;22(8):1085–1095. doi:10.1038/mp.2017.61

47. Haroon E, Raison CL, Miller AH. Psychoneuroimmunology meets neuropsychopharmacology: translational implications of the impact of inflammation on behavior. Neuropsychopharmacology. 2012;37(1):137–162. doi:10.1038/npp.2011.205

48. Gkesoglou T, Bargiota SI, Iordanidou E, Vasiliadis M, Bozikas VP, Agorastos A. Prognostic significance of blood-based baseline biomarkers in treatment-resistant depression: a literature review of available studies on treatment response. Brain Sci. 2022;12(7):940. doi:10.3390/brainsci12070940

49. Mayberg HS, Brannan SK, Tekell JL, Silva JA, Mahurin RK, McGinnis S, Jerabek PA. Regional metabolic effects of fluoxetine in major depression: serial changes and relationship to clinical response. Biol Psychiatry. 2000;48(8):830–843. doi:10.1016/s0006-3223(00)01036-2

50. Song EJ, Tozzi L, Williams LM. Brain circuit-derived biotypes for treatment selection in mood disorders: a critical review and illustration of a functional neuroimaging tool for clinical translation. Biol Psychiatry. 2024;96(7):552–563. doi:10.1016/j.biopsych.2024.03.016

51. Etkin A. A reckoning and research agenda for neuroimaging in psychiatry. Am J Psychiatry. 2019;176(7):507–511. doi:10.1176/appi.ajp.2019.19050521

52. Woo CW, Chang LJ, Lindquist MA, Wager TD. Building better biomarkers: brain models in translational neuroimaging. Nat Neurosci. 2017;20(3):365–377. doi:10.1038/nn.4478

53. Aloezos C, Wai JM, Bluth MH, Forman H. Use of the clinical laboratory in psychiatric practice. Clin Lab Med. 2016;36(4):777–793. doi:10.1016/j.cll.2016.07.004

54. Dong Z, Shen X, Hao Y, Li J, Xu H, Yin L, Kuang W. Gut microbiome: a potential indicator for predicting treatment outcomes in major depressive disorder. Front Neurosci. 2022;16:813075. doi:10.3389/fnins.2022.813075

55. Diniz BS, Mulsant BH, Reynolds CF 3rd, Blumberger DM, Karp JF, Butters MA, et al. Association of molecular senescence markers in late-life depression with clinical characteristics and treatment outcome. JAMA Netw Open. 2022;5(6):e2219678. doi:10.1001/jamanetworkopen.2022.19678

56. Greden JF, Parikh SV, Rothschild AJ, Thase ME, Dunlop BW, DeBattista C, et al. Impact of pharmacogenomics on clinical outcomes in major depressive disorder in the GUIDED trial: a large, patient-and rater-blinded, randomized, controlled study. J Psychiatr Res. 2019;111:59–67. doi:10.1016/j.jpsychires.2019.01.003

57. Oslin DW, Lynch KG, Shih MC, Ingram EP, Wray LO, Chapman SR, et al. Effect of pharmacogenomic testing for drug-gene interactions on medication selection and remission of symptoms in major depressive disorder: the PRIME Care randomized clinical trial. JAMA. 2022;328(2):151–161. doi:10.1001/jama.2022.9805

58. Bousman CA, Arandjelovic K, Mancuso SG, Eyre HA, Dunlop BW. Pharmacogenetic tests and depressive symptom remission: a meta-analysis of randomised controlled trials. Pharmacogenomics. 2019;20(1):37–47. doi:10.2217/pgs-2018-0142

