## Supplemental material for "Neuroplasticity-Based Treatment Prioritization in Major Depressive Disorder: A Two-Cohort Study"

**Supplementary Material**: **Neuroplasticity-Based Treatment Prioritization in Major Depressive Disorder**

Supplementary statistical detail

Standardization. NPS was standardized within cohort in two ways: as a percentile (0–100) for banding and display, and as a z-score for regression models, with effects reported per 1-SD increase. Both transformations are strictly monotone within cohort and therefore leave all rank-based statistics, including the area under the receiver operating characteristic curve (AUC), unchanged. Because the two cohorts were assayed on different absolute scales, no analysis pools raw NPS across cohorts.

Uncertainty. Confidence intervals for proportions are exact binomial (Clopper–Pearson). Confidence intervals for the AUC and confidence bands for the logistic response-probability curves were obtained by resampling patients (not episodes) with replacement, 500 iterations, so that uncertainty reflects the number of independent participants rather than the number of episodes.

Receiver operating characteristic curves. With 40 and 35 episodes, the empirical ROC is a coarse step function. For display only, the curves in Figure 4 of the main text were smoothed by estimating the score densities in responding and non-responding episodes with a Gaussian kernel (bandwidth 0.3) and integrating them. All reported AUC values are computed from the empirical data, not from the smoothed curves. A binormal parameterization was considered and rejected: the Cohort 1 score distribution is right-skewed (skewness 1.75) and the binormal fit inflated the AUC to 0.998.

Supplementary methods

**iPSC reprogramming.** iPSCs were reprogrammed from PBMCs for the BDNF studies and for all samples collected from Sheba and Geha Medical Centers. iPSCs were reprogrammed from LCLs for the STAR*D cohort. Reprogramming was carried out using the CytoTune-iPS 2.0 Sendai reprogramming kit (Thermo Fisher). Briefly, 2.5 × 10^5^ cells were resuspended in 1 ml of SeV-supplemented with media, centrifuged at 1000 × g for 30 min and incubated for 24 h at 37 °C in a single well of 12-well plate. Media was replaced next day and virus particles washed. Two days later, cells were transferred to (hESC)-qualified Matrigel (Corning) coated six-well plates. Media was gradually transitioned to ReproTeSR (STEMCELL Technologies) then switched to mTeSRPlus (STEMCELL Technologies) at 25% confluency. Cells were passaged at 1:6-1:10 ratios upon reaching 70-80% confluency and were mycoplasma free (AssayGenie).

**Cortical neuron differentiation.** Cortical neuronal differentiation was carried out as previously described with modifications (main text reference 30). iPSCs were transduced with a TetO-inducible NGN2 lentiviral vector with puromycin resistance and selected with puromycin. Cells were seeded on hESC-qualified Matrigel–coated 96-well plates at 3 × 10³-1 × 10⁴ cells/well in NGN2 medium 1 (Table S1). The next day, two 50% medium changes were performed with ROCK inhibitor–free medium 1. On day 2, two 50% medium 2 changes were done (Table S1). 50% media changes were done every other day with media 2 until d10, and from d11 to d14 with media 3 (Table S1). All selected patient-derived LCLs/PBMCs were successfully reprogrammed into iPSCs and differentiated into cultures enriched for excitatory cortical neurons.

**Table S2. Compound concentrations.**

| **Compound** | **2× medium (µM)** | **Final on cells (µM)** |
| --- | --- | --- |
| Sertraline | 0.5 | 0.25 |
| Citalopram | 0.25 | 0.125 |
| Escitalopram | 2.5 | 1.25 |
| Paroxetine | 0.5 | 0.25 |
| Venlafaxine | 0.25 | 0.125 |
| Bupropion | 0.25 | 0.125 |
| Nortriptyline | 0.125 | 0.0625 |
| Mirtazapine | 0.025 | 0.0125 |
| Quetiapine | 1.25 | 0.625 |

Compounds were prepared at 2× in medium and applied as a 50% medium exchange on days 13, 15 and 17 of differentiation, giving the final on-cell concentrations shown. Vehicle was DMSO.

**Table S1. Differentiation medium protocol.**

| **Neuronal Induction (NI) media 1 (Day 0-1)** | **50 ml** | **Final concentration** |
| --- | --- | --- |
| DMEM:F12 | 49 ml |  |
| NEAA | 500 μl |  |
| N2 | 500 μl |  |
| BDNF | 2 μl | 5 ng/ml |
| NT-3 | 10 μl | 5 ng/ml |
| Laminin |  | 0.2 μg/ml |
| Doxycycline | 10 μl | 2 μg/ml |

| **Neuronal Induction (NI) media 2 (Day 2-10)** | **50 ml** | **Final concentration** |
| --- | --- | --- |
| Neurobasal medium | 48 ml |  |
| B27 | 1 ml |  |
| Glutamax | 500 μl |  |
| P/S | 500 μl |  |
| BDNF | 2 μl | 5 ng/ml |
| NT-3 | 10 μl | 5 ng/ml |
| Laminin |  | 0.2 μg/ml |
| Doxycycline | 10 μl | 2 μg/ml |

| **Neuronal Induction (NI) media 3 (Day 11 onwards)** | **50 ml** | **Final concentration** |
| --- | --- | --- |
| Neurobasal medium | 48 ml |  |
| B27 | 1 ml |  |
| Glutamax | 500 μl |  |
| P/S | 500 μl |  |
| BDNF | 2 μl | 5 ng/ml |
| NT-3 | 10 μl | 5 ng/ml |
| Laminin |  | 0.2 μg/ml |

Figure S1. **Characterization of iPSCs and their differentiation into cortical neurons.**
 **(A)** Representative phase-contrast image showing typical iPSC colony morphology. **(B)** Flow cytometry analysis of pluripotency markers SSEA4 and TRA-1-60, demonstrating a high proportion of double-positive iPSCs (n=20). **(C)** Quantification of SSEA4-positive cells across 20 independent reprogrammed iPSC lines, showing consistently high pluripotency. **(D)** Representative immunofluorescence image of differentiated cortical neurons stained for Bassoon (red) and Homer (green), with GFP-labeled neurons (green), indicating synaptic protein localization along neuronal processes. Scale bar: 20 µm. **(E)** Flow cytometry analysis of CTIP2 expression during neuronal differentiation at day 8, day 14, and day 21, indicating progressive enrichment of CTIP2-positive cortical neurons over time. **(F)** qRT-PCR analysis showing fold-change expression of the neuron markers **VGLUT1** and **VGLUT2** in differentiated neurons compared with the iPSC pool. Data are shown as mean ± SEM; ***p < 0.001 (n=3). **(G)** Quantification of Syn1-positive cells across 20 differentiated neuronal lines (n=20).

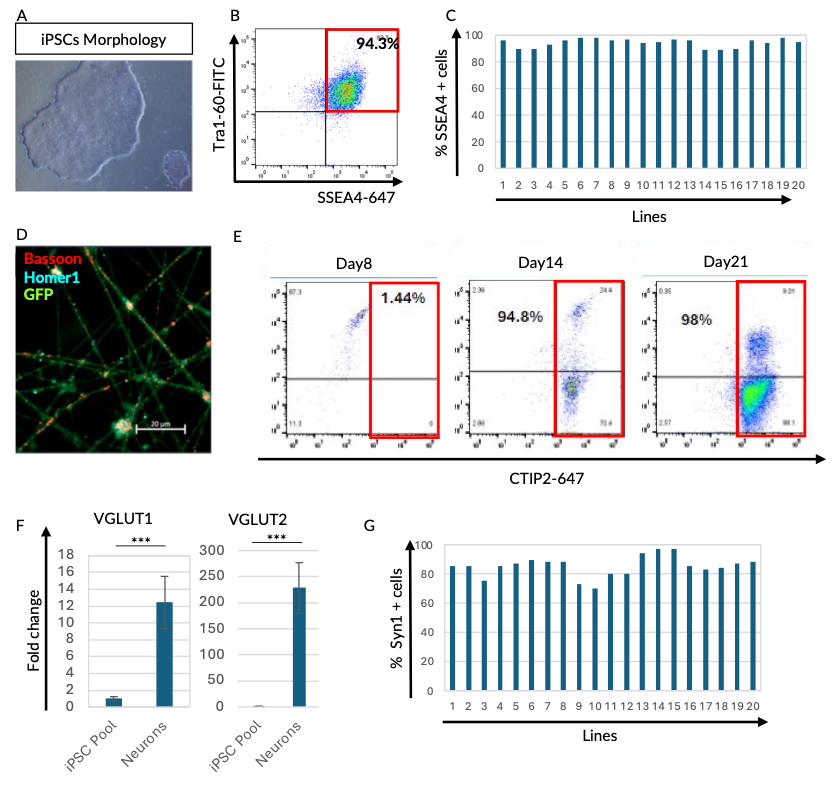

Figure S2. **Representative brightfield images showing morphology of iPSC-derived cortical neurons and technical reproducibility of Neuroplasticity Score (NPS).**

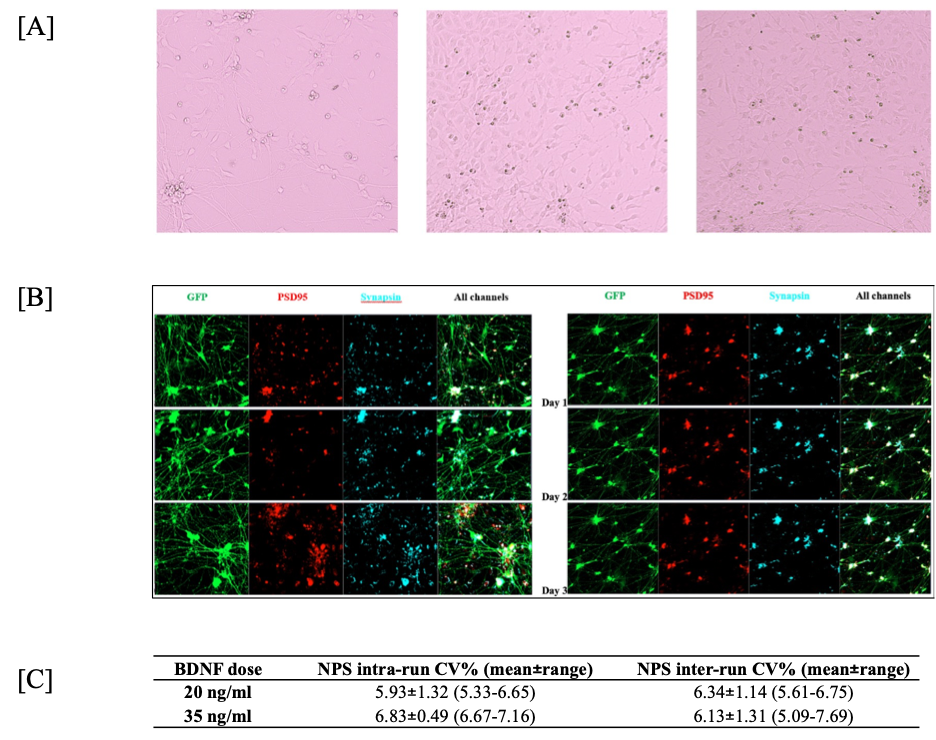

 [A] Brightfield images demonstrating neuronal morphology in iPSC-derived cortical neurons generated from peripheral blood samples. Images are representative of multiple donors and differentiation batches. [B] Representative regions of interest from repeated immunostaining and imaging runs across operators and days, demonstrating consistent marker intensity and neuronal morphology. [C] Reproducibility of NPS across independent differentiations and staining runs from three donors at two BDNF concentrations (20 and 35 ng/mL), showing coefficient of variation <10% across replicates.

**Figure S3. Calibration of NPS-based response probability.**

Observed against predicted response rate within each NPS tertile, for each cohort. Predicted rates are the mean fitted probability from the cohort-specific logistic model; observed rates are the proportion of responding episodes in the same tertile. The dashed line is perfect calibration. Cohort 1: predicted 18%, 49% and 81% against observed 23% (3/13), 38% (5/13) and 86% (12/14). Cohort 2: predicted 14%, 43% and 78% against observed 9% (1/11), 42% (5/12) and 83% (10/12).

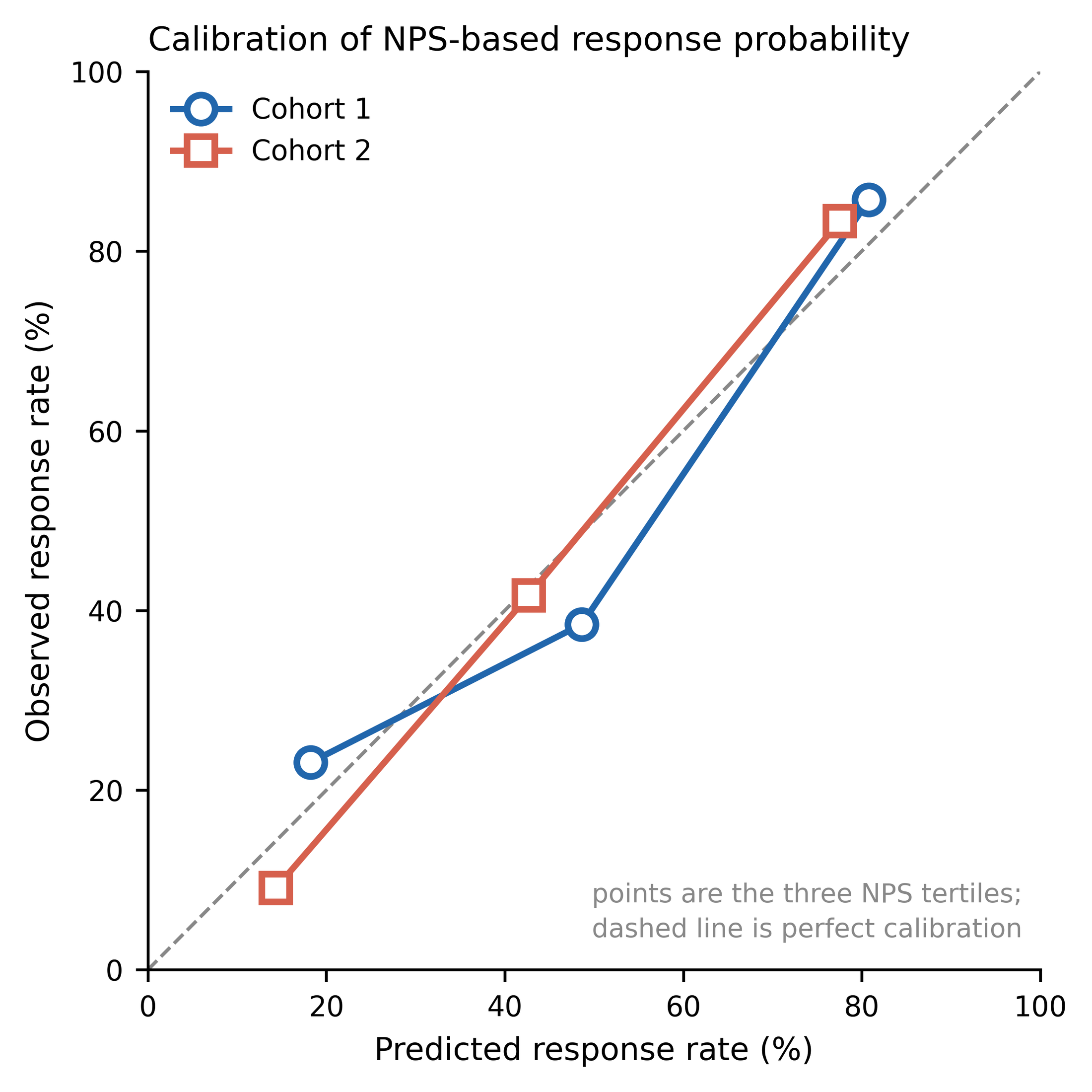

**Figure S4. Per-episode within-patient detail.**

Every treatment episode for each of the 22 patients who contributed both a responding and a non-responding episode (the patients summarized in Figure 5 of the main text). Each row is one patient; each point is one treatment episode, positioned by its NPS percentile within cohort and coloured by whether that episode met response criteria. The ringed point is the patient's highest-scoring drug, and the label at right records whether that drug was one the patient responded to. Patients are numbered separately within each cohort.

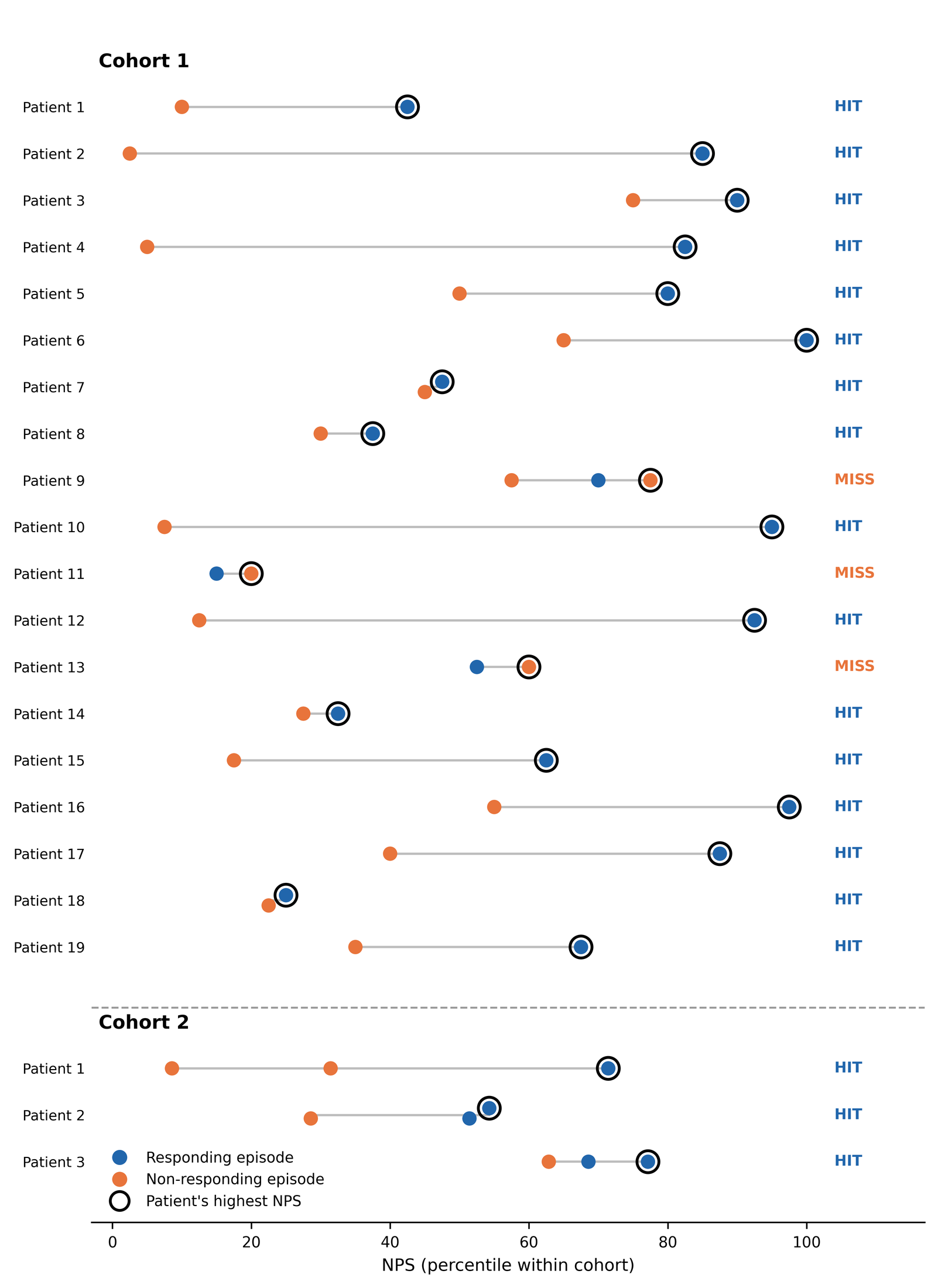
